# LITERATURE REVIEW ON INTERVENTIONS FOR PREVENTION OF UNPLANNED EXTUBATION IN NICU PICU: A PHILOSOPHY OF NURSING SCIENCE

**DOI:** 10.1101/2023.10.05.23296414

**Authors:** Mariyam Mariyam, Moses Glorino Rumambo Pandin, Netty Prasetiya Fitriani

## Abstract

**Introduction:** One of the complications of endotracheal tube (ETT) installation is unplanned extubation (UE). UE is an initial inappropriate removal of an ETT by a patient or medical personnel. This literature review are to describe interventions to prevent unplanned extubation in NICU and PICU.

**Method:** Literature review searches using Sciencedirect, Google Scholar and Pubmed. The keywords used in searching articles were unplanned extubation PICU NICU. The search criteria were publication year 2017-2023, quasi-experimental design and full text.

**Result:** The results show that actions that can reduce the incidence of UE are standardization interventions for ETT safety, protocols for moving patients (patient transportation), and collaborative interventions for providing sedation.

**Conclusion:** standardization of ETT safety, protocols for moving patients (patient transport), and collaboration in providing sedation can be used to reduce unplanned extubation.

## Introduction

Respiratory failure is an indication that a child should be treated in the NICU or PICU. One of the measures taken to support a child’s breathing is the installation of an endotracheal tube (ETT). During ETT installation, one of the complications is unplanned extubation (UE) (Mahaseth et al., 2020).

UE is the unintentional disengagement of an ETT by a patient or healthcare worker (Crezeé et al., 2018)(Mahaseth et al., 2020). Causes of UE include oral grooming, turning and patting the back, restraints, and airway care (Wang et al., 2021)

The impact of UE on children treated in the NICU and PICU is around 20% have cardiovascular collapse and require cardiopulmonary resuscitation (Kandil et al., 2018). Most of the cases (91.7%) of EU are taken seriously accidents that have the potential to result in arrhythmia and aspiration (Chao et al., 2017).

The results of previous research show 137 patients with ETT installed, 44 patients experienced UE (Cho & Yeo, 2022) and of 1,721 patients with invasive ventilation, 39 patients experienced UE (UE rate 2.27%)(Kodicherla et al., 2021).

Interventions carried out to reduce UE include ETT safety standardization, protocols for moving patients (patient transportation), and collaboration in providing sedation (Nelson et al., 2022, Obina & Almohileb, 2022). UE risk factors include the child’s birth weight, gestational age, consciousness status, duration of ventilator installation, ventilator weaning, duration of endotracheal intubation, ETT fixation, duration of ETT fixation, nurse to patient ratio and nurse workload, shifts (for example morning, afternoon or night shifts), and other nursing actions (suction, bed transfer, weight weighing, ETT re-fixation) (Cho & Yeo, 2022).

Children who experience UE will experience hypercarbia and hypoxia. Children require ongoing care and require ventilatory support through resuscitation, restoring airway patency, increasing types of care which results in increased costs of caring for children in the NICU and PICU, airway trauma, cardiac arrest and death, increased hemodynamic instability, infection and intraventricular hemorrhage (Crezeé et al., 2018, Lauderbaugh & Sutherland, 2020).

EU prevention efforts include implementing standard ETT safety procedures, maintaining vigilance regarding ETT installation so that it does not move position, staff education, maintaining an adequate nurse/patient ratio and implementing standard care practices and/or protocols (Minda et al., 2022) (Hu et al., 2017) monitor intubated patients, ETT management and restraint (Jayawardena et al., 2021). Healthcare professionals must identify cases at high risk of UE, identify causes of UE, and utilize appropriate interventions to prevent UE. Proper ETT securing methods, training in maintaining the ETT during the procedure, adequate nurse patient ratio, timely weaning of mechanical ventilation, and implementation of bundles for improving the quality of UE are some of the key points of reducing the incidence of UE in NICU PICU (Angurana, 2021).

The purpose of this literature review is to identify interventions to prevent unplanned extubation in NICU and PICU.

### Search

This research uses a literature review approach regarding interventions to prevent UE incidents. Inclusion criteria include publication time 2017-2023, quasi-experimental research type, original research article, full text available. Literature search used Google Scholar, Sciencedirect and Pubmed. The keywords used in this literature search are Unplanned extubation, NICU, PICU. The search results showed that there were 806 articles obtained according to keywords, then searched and adjusted to the inclusion criteria, 7 research articles were obtained..

**Fig 1.**
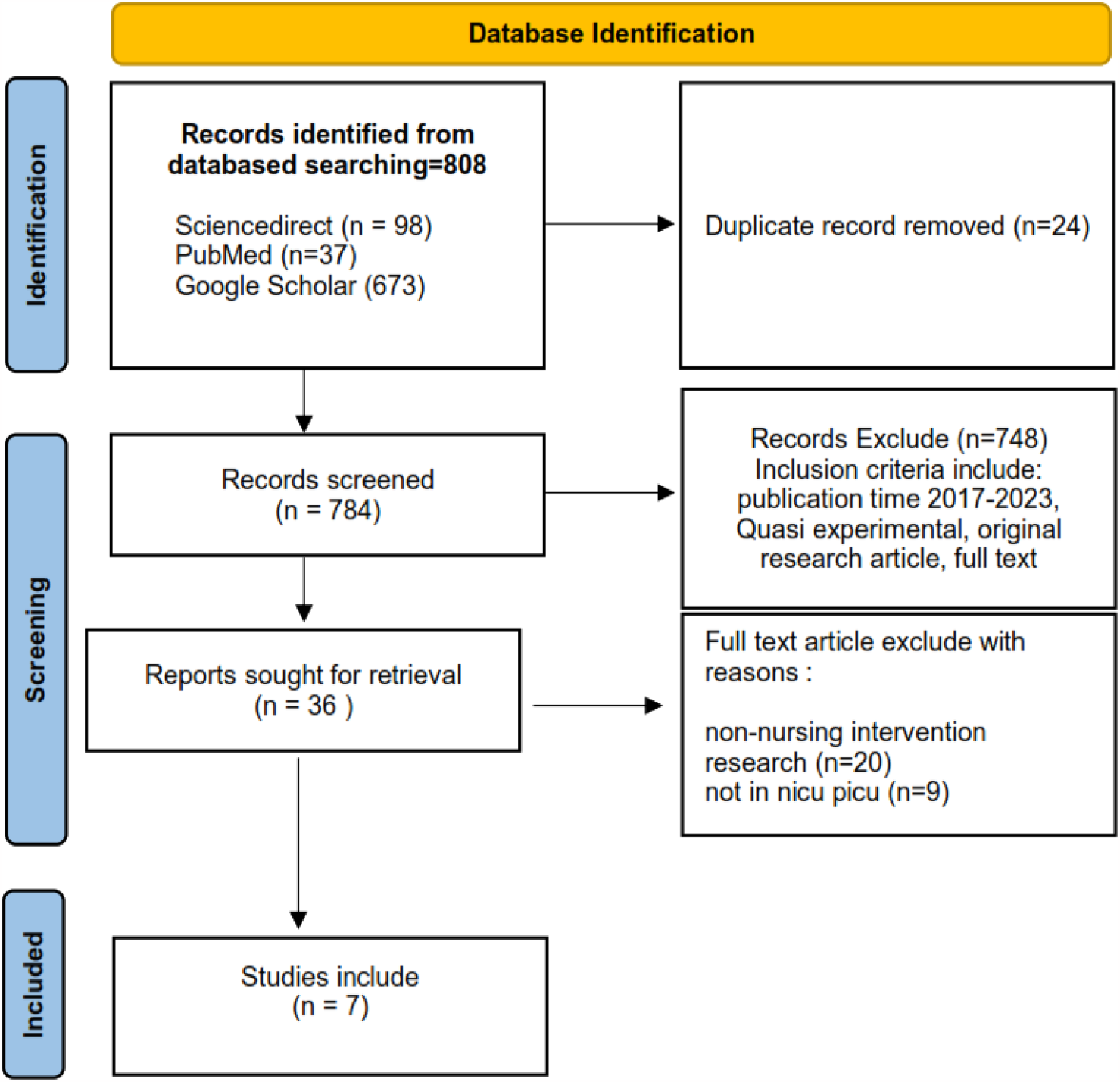
PRISMA Flowcart

**Table 1.**
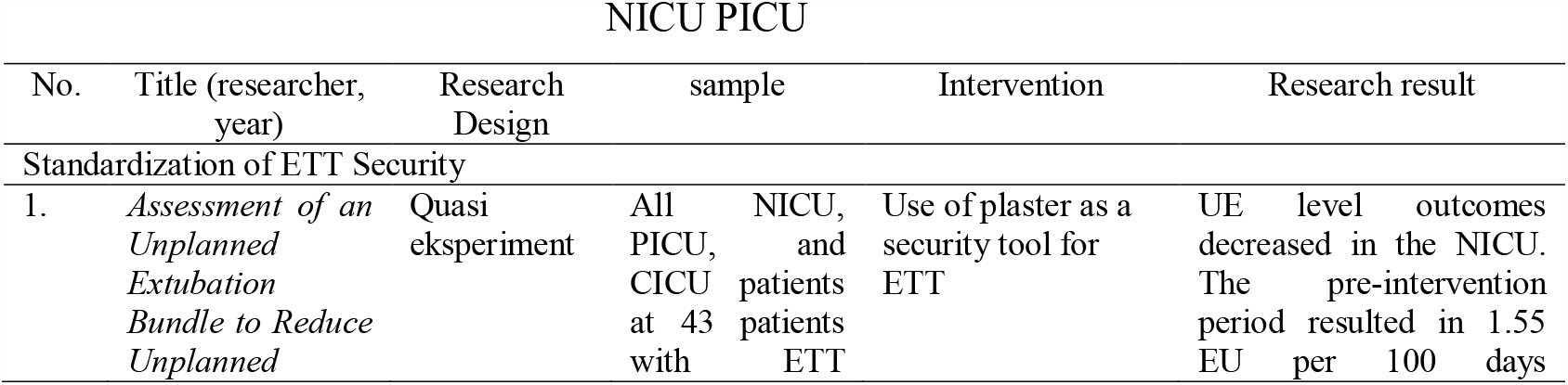
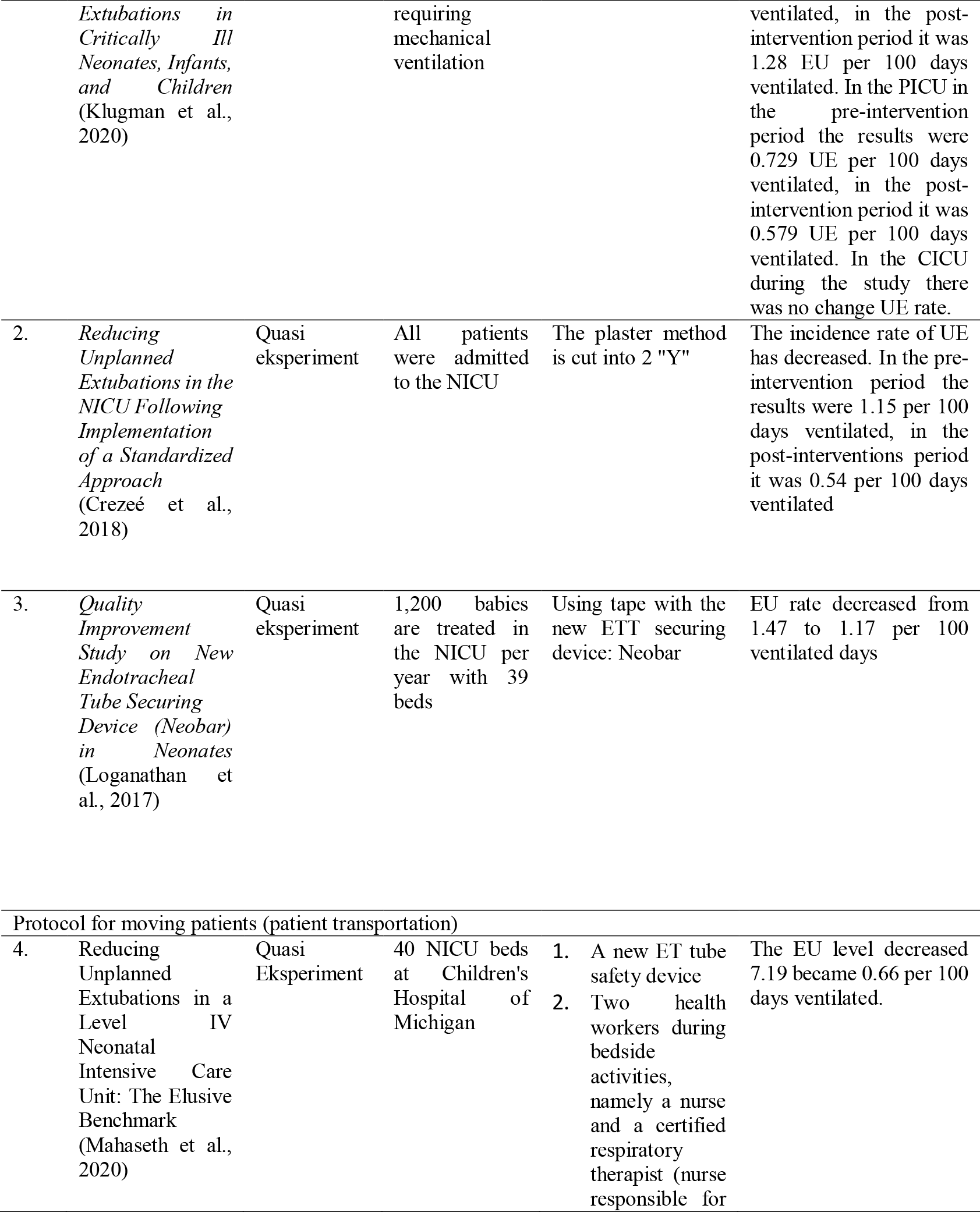
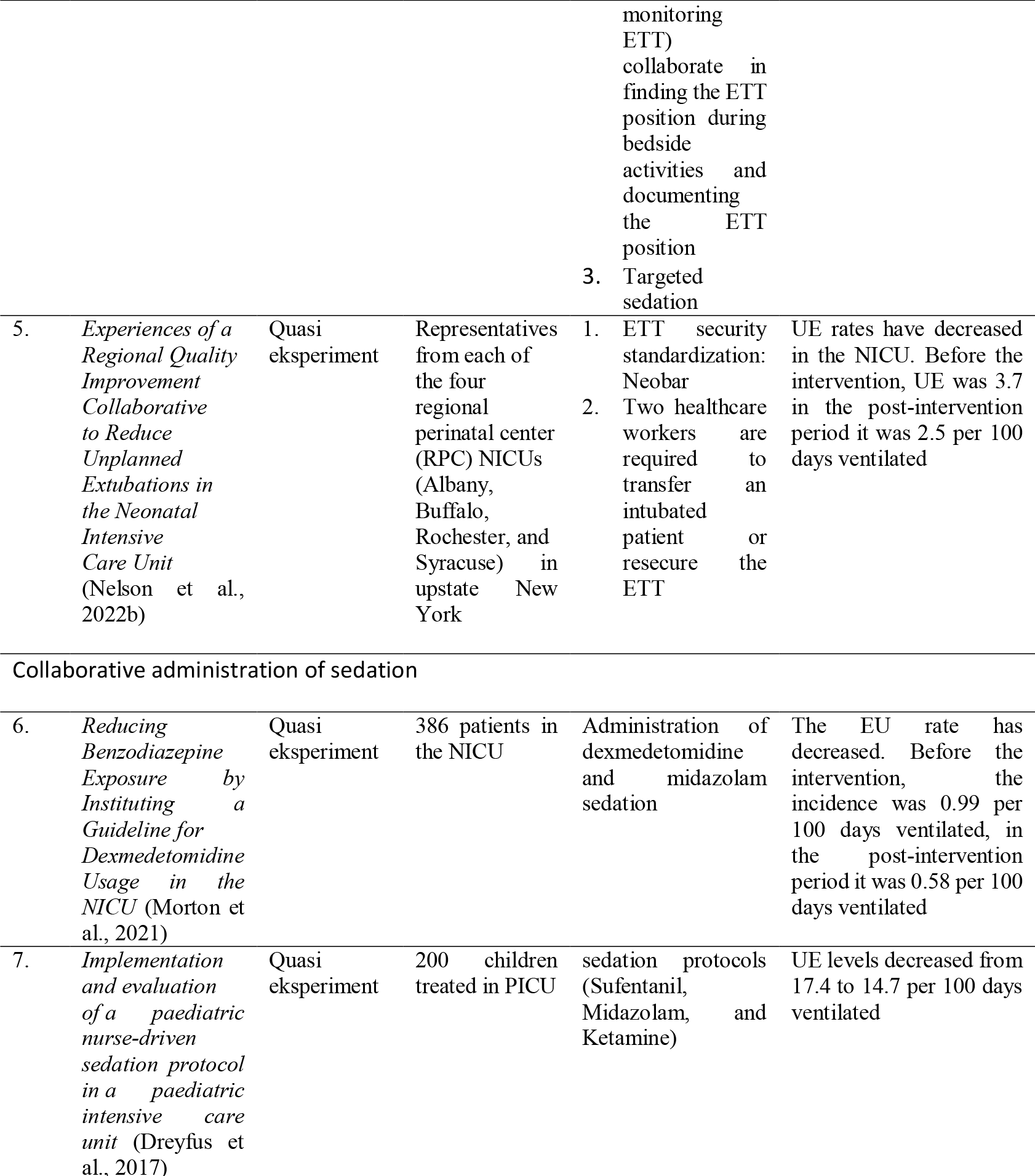
Results of Data Synthesis Interventions to Prevent Unplanned Extubation in NICU PICU.

## Results and Discussion

### 1. Standardize ETT security

The ETT safety standardization technique is a form of ETT safety standardization that poses the least risk of skin damage, is easy to implement and has the fewest UE incidents (Loganathan et al., 2017). ETT safety standardization interventions can be carried out using plaster, the plaster method is cut into two “Y” shapes, and the use of a new ETT security tool (Neobar).

The use of plaster for the safety of the ETT per unit is different, because premature babies have thinner skin and are still sensitive, the short length of their trachea means that the ET position is easily removed. Previous research also explained that the use of plaster as a safety measure for ETT had decreased the incidence of UE (Hatch et al., 2019). Health workers, including doctors, nurses or respiratory therapists, have the same role in resetting or repositioning the ETT when a UE event occurs. When the ETT position is incorrect, all health workers have the same role in maintaining the ETT position. Interventions to maintain the ETT position include if the loose plaster is put back on, if the ETT position is not suitable then it must be checked again, and whether oxygen can be administered or not, all are responsible for patient safety (Klugman et al., 2020).

ETT safety standardization intervention with the plaster method cut into 2 “Y” shapes can reduce the incidence of UE. How to use it: one is placed on the upper lip and the other is placed on top of the ET from one end of the corner of the mouth (Crezeé et al., 2018). The same study implemented standardization of ETT safety procedures using plaster with the plaster method being cut into 2 “Y” shapes and medical personnel always observing directly to ensure standardization of the safety of the ETT used, so that previous research could reduce the incidence of UE (Kandil et al., 2018).

The next intervention is related to the use of new models of ETT security tools such as Neobar which can reduce the incidence of UE. Neobar is an latex-free hydrocolloid at both ends to secure the ETT. The advantage of NeoBar compared to plaster is that Neobar has various sizes with different color codes, Neobar can be used on babies with very low birth weight, the ET size on Neobar is easier to see without having to remove the plaster and looks neater (Loganathan et al., 2017, Mahaseth et al., 2020). Other study also apply the safety of ETT using Neobar to reduce the incidence of UE in the NICU (Aydon et al., 2018)

### 2. Protocol for moving the patient (patient transportation)

Protocol intervention for moving patients (patient transportation) first with the use of a new ET tube safety device, namely Neobar, 2 health workers during bedside activities, namely a nurse and a certified respiratory therapist (nurse responsible for monitoring ETT) working together in searching for ETT positions, documentation of ETT positions and target sedation based on infant respiratory support, infant conditions that may decrease the incidence of UE. Nurses need to reconfirm the location of the ETT when the patient has been transferred to the NICU room (Mahaseth et al., 2020).

The second intervention protocol for moving patients (patient transport) standardizes ETT safety: Neobar and two health workers are required to move intubated patients or re-secure the ETT. The study used this collaborative intervention to prevent the incidence of UE so that each NICU could reduce the incidence of UE (Nelson et al., 2022)

Transferring a patient requires a minimum of 2 health workers consisting of nurses and other therapists to help identify and ensure that the ETT is in the right position during transportation. Nurses need to pay attention to the position of the ETT when carrying out several actions, including when providing interventions to patients, when carrying out radiography procedures, transporting patients or repositioning them. The nurse must pay attention to the location of the ETT so that it does not change position (Kandil et al., 2018). Documentation of ETT position with a grading scale (Nelson et al., 2022)

Predisposing factors for UE can be related to patients, health workers, activities, and shift time factors. Male patients are more at risk of experiencing UE than female. Men have stronger energy than women because there is the testosterone hormone in men which increases mass. This can result in the ETT dislodging itself. Patients treated by junior medical personnel are more at risk of experiencing UE compared to senior medical personnel, because junior medical personnel are still learning and have less skills in managing awake patients (Minda et al., 2022).

Patient activities can also experience UE events, because they feel uncomfortable and want to remove the ETT. The patient’s awareness is associated with the occurrence of UE, because when the patient is agitated he becomes unstable and moves a lot. Shift times can experience UE events, especially night shifts. Nurses on night shifts work longer hours than morning and afternoon shifts. Nurses who work the night shift may spend time during the day doing other work and become exhausted at night. Patients who sleep at night may be disturbed by the ventilator monitoring, causing patient to wake up, so that the nurse becomes sleepy because they have to meet the patient who is awake (Minda et al., 2022).

Patient monitoring during transport by medical personnel and equipment is a key of risk prevention. Faster transport times and a team of medical personnel performing specialized transport were associated with better outcomes, while severity illness was a sign of undesirable complications. The type of monitoring during transport varies greatly with the environment, medical personnel skills, and the severity of the patient’s illness (Branson & Rodriquez, 2020).

### 3. Collaborative administration of sedation

Installing an ETT on a child can cause physical discomfort to the child and trigger a stress response. Children who are mechanically ventilated will receive appropriate sedation (Morton et al., 2021,Wang et al., 2021)

Midazolam is currently a sedative commonly used for mechanical ventilation in children, but it has many disadvantages, because it is not effective in inhibiting the stress response caused by ETT (Wang et al., 2021). Midazolam can be administered orally, rectally, intramuscularly (IM), and intravascularly (IV). Side effects include dependence on midazolam (Egbuta & Mason, 2021)

Dexmedetomidine is an alpha-2 adrenergic agonist. Dexmedetomidine is an α2 adrenergic receptor agonist. This drug does not inhibit breathing and has a protective effect on the function of the heart, brain, kidneys and other organs (Wang et al., 2021). Dexmedetomidine can be given IV, IM and sublingually. The sedation effect of dexmedetomidine is very similar to natural sleep. Dexmedetomidine as a primary sedative is appropriate, safe for children and can reduce midazolam exposure. Side effects of using dexmedetomidine are hypotension and bradycardia. Dexmedetomidine ensures comfort, avoids the use of other sedatives, and reduces delirium (Egbuta & Mason, 2021)(Morton et al., 2021). Previous studies also compared dexmedetomidine with midazolam in adults, finding that dexmedetomidine can reduce the duration of mechanical ventilation and shorten the time to extubation (Egbuta & Mason, 2021).

Sedation protocols (sufentanil, midazolam, and ketamine) in collaboration with health workers where sedation to children is carried out by nurses in the PICU can reduce the incidence of UE (Dreyfus et al., 2017)

## Conclusion

The use of ETT in children treated in the NICU and PICU has the possibility of UE. EU preventive interventions include ETT safety standardization which can be carried out using plaster, the plaster method is cut into two “Y” shapes, and the use of a new ETT security tool, namely Neobar. The protocol for moving patients (patient transportation) involves health workers including doctors, nurses or respiratory therapists. Using the new ET tube safety device, 2 health workers during bedside activities, namely a certified nurse and respiratory therapist, collaborate in finding the ETT position during bedside activities and documenting the ETT position and collaboration in administering sedation.

## Data Availability

All data produced in the present work are contained in the manuscript

